# Efficacy and safety of convalescent plasma versus standard care in hospitalized patients with COVID-19 from the Peruvian Social Security Health System: open-label, randomized, controlled clinical trial

**DOI:** 10.1101/2022.09.21.22280195

**Authors:** Cristian Villanueva, Ibeth Neyra, Arturo Sagastegui, Ausberto Chunga, Martin Oyanguren, Martina Guillermo-Roman, Suly Soto-Ordoñez, Jorge L. Maguiña, Yamilee Hurtado-Roca, Percy Soto-Becerra, Roger V. Araujo-Castillo

**Author notes:** **Corresponding:** Roger V. Araujo-Castillo, MD, Address: Cápac Yupanqui 1400 - Jesus María, Lima 11, Perú. **Financing** This clinical trial has been co-financed by two institutions of the Government of Peru, the Institute for the Evaluation of Technologies in Health and Research - IETSI of the Social Security of Health (EsSalud) and the National Council of Science, Technology and Technological Innovation (CONCYTEC) through peer-reviewed external financing: the National Fund for Scientific Research, Technological Development and Technological Innovation (FONDECYT), subsidy (Nº 068-2020-FONDECYT). This grant was awarded to CV, IN, AS, AC, MO, JLM, YH, PSB and RA. The funders had no role in study design, data collection and analysis, decision to publish, or manuscript preparation. **Data availability** The raw data was generated at the Institute for the Evaluation of Technologies in Health and Research - IETSI of Peru’s Social Security of Health (EsSalud). Restrictions apply to the public availability of these data due to institutional patient data sharing policies. However, the data is available upon reasonable request from the author. **Clinical Trial Registration Number:** This clinical trial has been registered in the Peruvian Registry of Clinical Trials (REPEC, by Spanish acronym) with the following ID: PER-013-20.

## Abstract

**OBJECTIVES:** To assess the efficacy and safety of convalescent plasma plus standard of care (CP + SoC) compared with standard of care (SoC) alone in patients hospitalized for moderate to severe COVID-19 who do not yet require mechanical ventilation.

**METHODS:** Phase 2 randomized, parallel-group, randomized, open-label, controlled, superiority, single-center clinical trial. This clinical trial has been registered in REPEC with the following ID: 013-20. Hospitalized adult patients with moderate to severe COVID-19 were enrolled. The allocation ratio was 1:1 in a variable-size permuted block randomization scheme. The primary outcome was death 28 days after the intervention. Secondary outcomes were mortality at 14 and 56 days, time to death at 56 days, time in the ICU at 28 days, time on a mechanical ventilator at 28 days, frequency of adverse events, and frequency of serious adverse events.

**RESULTS:** A total of 64 participants were enrolled, 32 were assigned to CP + SoC, and 32 to SoC. One participant assigned to CP + SoC withdrew his informed consent before applying the treatment. At day 28, there were no statistically significant differences for the primary outcome between the CP + SoC and SoC groups (relative risk: 2.06; 95%CI 0.73 to 7.11; p = 0.190). No differences were found in the incidences of mortality at 56 days (hazard ratio: 2.21; 95%CI 0.66 to 7.33; p = 0.182), admission to the ICU at 28 days (sub-hazard ratio: 2.06; 95%CI 0.57 to 8.55; p = 0.250), admission to mechanical ventilation at 28 days (sub-hazard ratio: 2.19; 95%CI 0.57 to 8.51; p = 0.260). Estimates for days 14 were similar. No infusion-related adverse events were reported during the study. There were no statistically significant differences in the frequency of any adverse events (odds ratio: 2.74; 95%CI 0.90 to 9.10; p = 0.085) or the frequency of serious adverse events (odds ratio: 3.60; 95%CI 0.75 to 26.1; p = 0.75).

**CONCLUSIONS:** No evidence was found that CP had a significant effect in reducing 28-day mortality. There was also no evidence that the frequency of adverse events was higher in those who received CP + SoC than those who received only SoC.

## Introduction

The SARS-CoV-2 virus causes coronavirus disease 2019 (COVID-19), identified in Wuhan, China, in December 2019 and declared a pandemic on March 12, 2020, a few months after the first case was reported (1–3). In the absence of available treatments, clinical trials initially focused on evaluating the replacement of interventions with recognized efficacy for other infectious diseases, such as antiparasitic, antiviral, antiinflammatory drugs, anticoagulants, and convalescent plasma, among others (4).

For more than a century, convalescent plasma (CP) has been used in the treatment of various diseases of viral origin: severe acute respiratory syndrome (SARS), Eastern respiratory syndrome (MERS), avian influenza A (H5N1), Spanish flu A H1N1 pandemic, among others (5,6). Theoretically, antibodies in the plasma of recovered COVID-19 individuals would be passive immunization agents for the immune system of patients with active disease (7); however, empirical evidence about its efficacy on important outcomes was anecdotal, coming mainly from case series or observational studies (5).

Initially, the studies showed conflicting evidence, and even the available systematic reviews and meta-analyses did not find consistent results (8–12). Some systematic reviews and meta-analyses concluded that PC shows a potential reduction in mortality, although with statistically uncertain estimates (8–10). However, other systematic reviews and meta-analyses concluded that PC does not offer any benefit to adverse outcomes of COVID-19 (11,12), but the quality of the evidence reviewed was low. These inconsistent results showed the need for more controlled clinical trials to clarify the uncertainty about the efficacy of PC in the treatment of COVID-19.

This clinical trial was conducted in this context of uncertainty about the efficacy of PC. However, as scientific evidence accumulated, it became increasingly clear that PC was ineffective in treating patients hospitalized for COVID-19 (13). For this reason, this study was terminated early. Although the current consensus indicates that there is high certainty that treatment with PC is not effective in reducing outcomes of death, admission to the ICU, or mechanical ventilation (13–15), there are still some controversies about whether these studies evaluated the doses, appropriate application times (16–21) and uncertainties about their safety (13). As of May 20, 2021, 100 clinical trials on CP had been registered, but only under 33% had been published (13), so the publication of the findings will contribute to resolving the uncertainties associated with CP therapy.

This study reports the results of a clinical trial that aimed to evaluate the efficacy and safety of PC plus standard of care (SoC) compared to SoC alone in outcomes of patients hospitalized for COVID-19. The main hypothesis of this clinical trial was that convalescent plasma treatment in patients with moderate to severe COVID-19, who do not yet require a mechanical ventilator, is effective in reducing 28-day mortality. Efficacy against intensive care unit (ICU) admission, ventilator, and adverse events were also evaluated. The article was written following the CONSORT 2010 guidelines (Consolidated Standards of Reporting Trials) (22).

## Methods

### Study design

Phase 2, randomized, controlled, open-label, parallel-group, superiority, single-center clinical trial. The study was approved by the Transitory National Research Ethics Committee (CNTEI) -COVID-19 through Certificate of Approval - CNTEI-007-2020 dated June 19, 2020. The study is registered in the Peruvian Registry of Clinical Trials (REPEC) with code PER-013-20 (23) and was approved by the National Institute of Health through Directorial Resolution 198-2020-OGITT-INS dated June 25, 2020. The last version of the approved protocol, translated to English for publication purposes, is available in S1 File. Ethical approval and informed consent form are in the S2 and S3 Files. A detailed description of procedures is available in the Manual of Procedures whose last version is in the S4 File. This study followed the CONSORT recommendations for reporting clinical trials (S5 File).

### Study population

This trial was conducted in the Emergency Service and the Transfusion Medicine Service of the Edgardo Rebagliati Martins National Hospital (HNERM), a tertiary care hospital located in Lima, the capital of Peru. Between September 2020 and April 2021, patients who met the following inclusion criteria were enrolled:

1. Adult male or female patient ≥18 years of age requiring hospitalization or hospitalized for COVID-19 without the need for mechanical ventilation (invasive or non-invasive) at the time of enrollment.
2. Written informed consent before performing study procedures.
3. Laboratory-confirmed diagnosis of SARS-CoV-2 infection by RT-PCR in nasopharyngeal or oropharyngeal swabs.
4. Patients at risk of progression of COVID-19 defined as the presence of two or more of the following laboratory values:
  a. Ferritin > 500 ng/mL
  b. D-dimer > 1 mg/L
  c. C-reactive protein > 15 mg/L
  d. Total lymphocytes <1000/mm3 or neutrophil/lymphocyte ratio >3.13
5. Or patients with a clinical manifestation of pulmonary compromise defined by the presence of two or more of the following clinical parameters
  a. Dyspnoea
  b. Respiratory rate greater than or equal to 30 per minute
  c. Oxygen saturation less than 93%
  d. PaO2/FiO2 less than 300 and pulmonary infiltrate greater than 50% in the 24 to 48 hours after the initial evaluation

Likewise, patients who met any of the following criteria were excluded:

1. Transfusion of any blood product within 120 days before administration of convalescent plasma.
2. Active pregnancy detected by a qualitative test that detects the hormone human chorionic gonadotropin (hCG) in the urine.
3. Current participation in a randomized clinical trial or past involvement in a clinical trial, and less than 30 days have passed since your last study visit.
4. Patient has life-threatening COVID-19 illness defined as one or more of the following:
  a. Respiratory failure, ventilatory type, defined as the need for invasive mechanical ventilation (with endotracheal intubation) or ECMO (extracorporeal oxygenation).
  b. Septic shock, defined as having criteria for sepsis (an increase of two or more points on the Sequential Organ Failure Assessment (SOFA) scale) (17) and requiring vasopressors to maintain MAP ≥ 65 mmHg after adequate hydration.
  c. Multiple organ dysfunction or failure, defined as the dysfunction of two or more systems other than the respiratory system. System dysfunction will be considered when a score of 2 or more is obtained on the SOFA scale in the following systems: coagulation, liver, cardiovascular, central nervous system, or kidney. The SOFA criteria used in this clinical trial are in the S1 Table.

### Study intervention

All participants received SoC for COVID-19. In addition, the treatment arm received ABO blood group system-compatible convalescent plasma from recovered COVID-19 patients (called donors) as an add-on therapy to the SoC. Other compatibilities, such as the Rh factor, were unnecessary for the plasma transfusion since it is free of red blood cells. Once a patient was assigned to the CP treatment arm, the CP bag was thawed, stored at 2-6°C, and used within 24 hours. A complete unit of plasma was administered intravenously as one dose, with a volume between 200-400 mL of convalescent plasma contained in a transfusion bag, at a recommended flow rate of 150-200 mL/h or less depending on patient tolerance. The plasma transfusion was in charge of one health personnel from the Transfusion Medicine Service who fulfilled the role of transfuser and was not part of the research team. The control arm received only SoC for COVID-19.

### Outcomes

The study’s primary outcome was the cumulative incidence of mortality (all causes) through day 28 after CP administration. Secondary outcomes were:

- Cumulative incidence of ICU admission at 14 and 28 days.
- Cumulative incidence of mechanical ventilation or extracorporeal oxygenation (ECMO) on day 14 and day 28 after randomization.
- Cumulative incidence of mortality (all causes) on days 14 and 56 after CP administration.
- Safety evaluations of CP + SoC compared to SoC alone up to day 28 considering the cumulative incidence of serious adverse events (SAEs) and infusion-related adverse reactions.

### Sample size

For an open-label, parallel-group, standard-of-care, controlled, randomized (1:1 ratio) superiority clinical trial and cumulative incidence of all-cause mortality at day 28 as the primary outcome, a sample size of 190 patients (95 per arm) assuming 21% mortality in the SoC arm (18) and an absolute difference of ∼14% (relative risk of 0.33 or ∼7% mortality in the CP arm), with a power statistic of 80% and a two-sided alpha level of 5% for a chi-square test of homogeneity without continuity correction. In addition, he estimated that approximately 63 PC donors would be needed.

### Procedures

The patients with COVID-19 were recruited at the HNERM Emergency Department through daily screening of medical records or on-site identification of the patients. Donors were invited through local print and audiovisual media advertising, which the Ethics Committee previously approved. Potentially eligible candidates were invited for a complete evaluation at the Blood Bank of the HNERM Transfusion Medicine Service.

The investigators of this study, certified and trained in Good Clinical Practices and Ethics in Research in Humans, conducted the process of obtaining the subject’s informed consent in accordance with Peruvian regulations and internationally accepted standards.

The patients received a presentation with key aspects of the clinical trial, they read the written informed consent document together with the investigator and their doubts were answered by him. In the end, the researcher confirmed that the information provided in the consent has been understood. When there were no more questions and the patient expressed understanding of the informed consent document, they were asked if she wishes to participate in the study. If accepted, the informed consent form was signed in duplicate by the patient or her legal representative, in case the patient is incapacitated, and by one of the researchers. In case she did not want to sign but did consent, her fingerprint was placed. Finally, one original informed consent form was delivered to the patient, and the other original was filed in a safe place. When the condition and severity of the patients who cannot consent did not allow the taking of informed consent in writing, consent was taken orally, recording the process in audiovisual media or digital images; and then, when feasible, obtaining the signature of the research subject in the written informed consent format. Due to the impediment to receiving medical visits that the COVID-19 services have imposed on the relatives of hospitalized patients, it was possible to contact legal representatives or relatives by phone or instant messaging to request their support or consent if the participant is prevented from doing so. Donors also received information about the clinical trial and gave their written informed consent before donating convalescent plasma. Patients and donors were informed about the possibility of collecting and storing an additional serum and plasma sample for up to one year for future use in research related to SARS-CoV-2. If they accepted, the participant or her legal representative signed written informed consent for future use of the biological sample.

Participants were randomly assigned to SoC alone (control arm) or treatment group (CP + SoC) with a 1:1 allocation according to a computer random number generator program that used permuted blocks of random size to ensure the balance of arms and the unpredictability of treatment assignments at any time during the trial. The random sequence was generated using the ado ralloc package (19) in Stata/SE version 16.1 for Microsoft Windows Pro 10 (StataCorp. 2019. College Station, TX: StataCorp LLC.).

To ensure concealment, block sizes were not disclosed until endpoint analysis and a central randomization scheme were implemented. The random assignment list was generated by a randomization officer and was kept hidden without sharing with any research team member until the clinical trial was completed. The randomization officer was a member of the team who was not part of the staff of evaluators or therapists, so integrity was guaranteed during the randomization process. A detailed timeline is provided in S1 Fig.

### Statistical analysis

The primary outcome was the cumulative incidence of death at 28 days after randomization. This analysis was by intention to treat. The effect of CP + SoC versus SoC alone on the cumulative incidence of mortality at 28 days was estimated using an adjusted relative risk (aRR) obtained from a log-binomial regression model that included the treatment variable and the block variable. Estimating the effect on mortality at 14 days followed the same approach described. However, the effect on 56-day mortality was assessed using a Cox regression that included treatment and block factor as covariates. The effect of CP + SoC versus SoC alone on these outcomes was estimated using adjusted hazard rate (HR) ratios. Survival curves were calculated using the Kaplan-Meier method and compared using the log-rank test. The effect of CP + SoC on admission to the ICU (at 14 and 28 days) and admission to mechanical ventilation (at 14 and 28 days), compared to SoC alone, was estimated using the sub-hazard ratio (subHR) considering death as a competitive event and obtained from a Fine and Gray model. Cumulative incidence functions were estimated and compared using Gray test. All analyzes were estimated with a 95% confidence interval and a significance level of 5%. Statistical analyzes were performed with R version 4.1.3 software.

## Results

### Patients

Between September 2020 and April 2021, 64 research subjects who met the selection criteria were enrolled, randomly assigning 32 to each study arm; One participant randomized to the intervention arm withdrew from the study before the application of PC, so 31 patients were assigned to convalescent plasma plus standard treatment and 32 to standard treatment alone (Fig 1).

**Fig 1.**
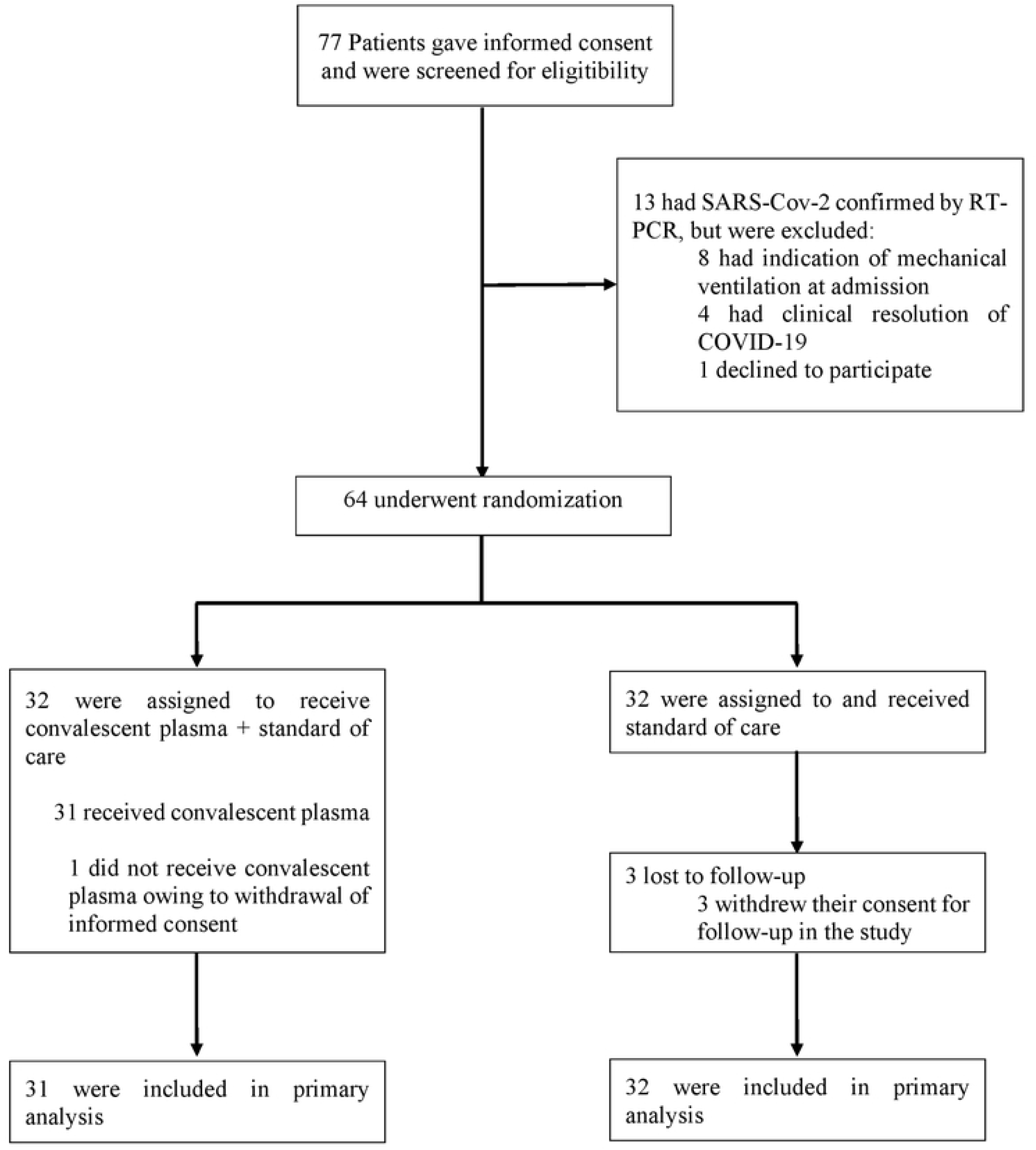
Enrollment and random assignment

The mean age of the patient population was 59.5 years (IQR: 46 to 72); 20.0% were women, and 20% had at least one comorbidity at study entry. The median time from onset of COVID-19 symptoms to enrollment was 13 days. The distribution of sociodemographic and clinical characteristics is shown in Table 1.

**Table 1.** Characteristics of the participants at enrollment

| Characteristics | CP + SoC<br>(n = 32) | Only SoC<br>(n = 32) |
| --- | --- | --- |
| **Sex** |  |  |
| Male | 23 (71.9%) | 28 (87.5%) |
| Female | 9 (28.1%) | 4 (12.5%) |
| **Age, years** | 62.5 (51.8, 72.0) | 56.5 (46.0, 69.0) |
| **Arterial hypertension** | 6 (18.8%) | 6 (18.8%) |
| **Mellitus diabetes** | 4 (12.5%) | 6 (18.8%) |
| **Pulmonary fibrosis** | 0 (0.0%) | 1 (3.1%) |
| **Asthma** | 0 (0.0%) | 1 (3.1%) |
| **Heart disease** | 1 (3.1%) | 1 (3.1%) |
| **Cerebrovascular disease** | 0 (0.0%) | 1 (3.1%) |
| **Obesity** | 9 (28.1%) | 7 (21.9%) |
| **Cancer** | 1 (3.1%) | 1 (3.1%) |
| **Hypothyroidism** | 0 (0.0%) | 2 (6.2%) |
| **Hemoglobin, g/dL** | 14.0 (13.2, 15.3) | 14.4 (13.2, 15.3) |
| **Hemoglobin categories** |  |  |
| <14 g/dL | 14 (45.2%) | 14 (43.8%) |
| 14-18 g/dL | 17 (54.8%) | 17 (53.1%) |
| >=18 g/dL | 0 (0.0%) | 1 (3.1%) |
| **Lymphocyte count, 1/uL** | 760.0 (625.0, 1,060.0) | 890.0 (737.5, 1,542.5) |
| **Hemoglobin categories** |  |  |
| <900/uL | 19 (61.3%) | 16 (50.0%) |
| 900-5200/uL | 12 (38.7%) | 16 (50.0%) |
| **Neutrophil count, 1/uL** | 9,360.0 (6,670.0, 12,845.0) | 7,520.0 (4,495.0, 9,385.0) |
| **Neutrophil Count Categories** |  |  |
| 1800-8000/uL | 12 (38.7%) | 17 (53.1%) |
| >8000/uL | 19 (61.3%) | 15 (46.9%) |
| **Platelet count, 1000/uL** | 318.0 (214.0, 420.5) | 262.5 (179.5, 380.0) |
| **Platelet Count Categories** |  |  |
| <130 x 1000/uL | 2 (6.5%) | 2 (6.2%) |
| 130-400 x 1000/uL | 19 (61.3%) | 23 (71.9%) |
| >400 x 1000/uL | 10 (32.3%) | 7 (21.9%) |
| **Prothrombin time, sec** | 11.1 (10.6, 11.9) | 11.0 (10.6, 11.9) |
| **PT Categories** |  |  |
| <10.5 seg | 6 (20.0%) | 5 (15.6%) |
| 10.5-13.0 seg | 22 (73.3%) | 25 (78.1%) |
| >13.0 seg | 2 (6.7%) | 2 (6.2%) |
| **Partial thromboplastin time, sec** | 36.1 (32.2, 39.6) | 34.0 (31.7, 36.4) |
| **TPT Categories** |  |  |
| 24.0-37.0 seg | 0 (0.0%) | 0 (0.0%) |
| >37.0 seg | 12 (100.0%) | 7 (100.0%) |
| **Serum glucose, mg/dL** | 136.0 (118.0, 183.0) | 137.0 (99.8, 185.5) |
| **Serum glucose categories** |  |  |
| 74-106 mg/dL | 3 (9.7%) | 10 (31.2%) |
| >106 mg/dL | 28 (90.3%) | 22 (68.8%) |
| **Serum creatinine, mg/dL** | 0.7 (0.6, 0.8) | 0.8 (0.7, 0.9) |
| **Glutamic-oxalacetic transaminase, U/L** | 50.0 (39.0, 82.5) | 58.5 (41.0, 67.5) |
| **TGO Categories** |  |  |
| 0.0-34.9 U/L | 0 (0.0%) | 0 (0.0%) |
| >34.0 U/L | 25 (100.0%) | 25 (100.0%) |
| **Glutamic-pyruvic transaminase, U/L** | 75.0 (48.0, 119.5) | 69.5 (51.2, 116.5) |
| **TGP Categories** |  |  |
| 0.0-49.0 U/L | 8 (25.8%) | 8 (25.0%) |
| >49.0 U/L | 23 (74.2%) | 24 (75.0%) |
| **Serum sodium, mmol/L** | 139.2 (136.8, 142.1) | 139.2 (136.8, 140.8) |
| **Serum sodium categories** |  |  |
| <132.0 mmol/L | 1 (3.2%) | 0 (0.0%) |
| 132.0-146.0 mmol/L | 30 (96.8%) | 32 (100.0%) |
| **Serum potassium, mmol/L** | 4.1 (3.9, 4.4) | 4.2 (4.0, 4.4) |
| **Serum potassium categories** |  |  |
| <3.5 mmol/L | 1 (3.3%) | 1 (3.1%) |
| 3.5-5.5 mmol/L | 29 (96.7%) | 31 (96.9%) |
| **C-Reactive Protein, mg/dL** | 9.2 (5.1, 15.4) | 6.1 (3.2, 11.2) |
| **PCR Categories** |  |  |
| 0.0-100.0 mg/dL | 31 (100.0%) | 31 (100.0%) |
| **Ferritin, ng/mL** | 822.0 (666.9, 1,446.0) | 869.9 (575.6, 1,362.5) |
| **Ferritin Categories** |  |  |
| 28.0-365.0 ng/dL | 3 (9.7%) | 4 (12.5%) |
| >365.0 ng/dL | 28 (90.3%) | 28 (87.5%) |
| **D-dimer, mg/mL** | 0.7 (0.4, 1.1) | 0.6 (0.5, 0.9) |
| **D-Dimer Category** |  |  |
| 0.00-0.54 ug/mL | 11 (39.3%) | 12 (40.0%) |
| >0.54 ug/mL | 17 (60.7%) | 18 (60.0%) |
| **Lactic dehydrogenase, U/L** | 390.0 (289.5, 479.0) | 347.5 (245.0, 403.8) |
| **DHL Categories** |  |  |
| 120.0-246.0 U/L | 5 (16.1%) | 9 (28.1%) |
| >246.0 U/L | 26 (83.9%) | 23 (71.9%) |
| **Quantification of anti-SARS-CoV-2 antibodies - IgG, AU/mL** | 39.5 (15.9, 69.1) | 37.8 (16.7, 67.5) |
| **Categories of Ab anti-SARS-CoV-2 IgG** |  |  |
| Non-reactive | 4 (12.9%) | 7 (21.9%) |
| Reactive | 27 (87.1%) | 25 (78.1%) |
| **Quantification of anti-SARS-CoV-2 antibodies - IgM, AU/mL** | 10.7 (2.3, 59.7) | 5.4 (2.0, 23.8) |
| **Categories of Ab anti-SARS-CoV-2 IgM** |  |  |
| Non-reactive | 15 (48.4%) | 20 (62.5%) |
| Reactive | 16 (51.6%) | 12 (37.5%) |
| **Pulmonary compromise by tomography** | 50.0 (40.0, 63.0) | 50.0 (42.0, 55.0) |

CP + SoC: Convalescent plasma plus standard of care; SoC: Standard of care alone.

### Primary outcome and secondary mortality outcomes

The 28-day mortality was 25.8% (8 of 26 patients) in the convalescent plasma plus standard therapy group and 12.5% (4 of 12 patients) in the standard therapy alone group. At day 28, although mortality in the CP + SoC group was twice that of SoC, these differences were not statistically significant (RR = 2.06; 95% CI 0.73 to 7.11; p = 0.190).

In the 56 days after enrollment, no statistically significant differences were found in the cumulative incidence curves of both groups (p = 0.196) (Fig 2). Similarly, there were no significant differences in the incidences of mortality (HR 2.21, 95% CI 0.66 to 7.33; p value = 0.182) (Table 2). The proportionality assumption of the Cox regression hazards was supported by the Grambsch and Therneau test (p = 0.450) and the Schoenfeld residual inspection.

**Table 2.** Clinical Results in patients who received CP + SoC compared with SoC only.

| Outcomes | Only SoC<br>(n = 32) | CP + SoC<br>(n = 31) | Risk Ratio or Hazard<br>Ratio<br>(95% CI); valor p |
| --- | --- | --- | --- |
| Primary outcome, death at 28 days;<br>No. events (%) | 4 (12) | 8 (26) | Risk ratio; 2.06<br>(0.73 a 7.11); 0.190 |
| Secondary outcomes |  |  |  |
| Death at 14 days; No. events (%) | 4 (12) | 7 (23) | Risk ratio; 2.06<br>(0.73 a 7.11); 0.190 |
| Time to ICU admission in 14<br>days; n events/person-time | 3 (384) | 6 (311) | Subhazard ratio; 2.21<br>(0.57 a 8.55); 0.250 |
| Time to ICU admission in 28<br>days; n events/person-time | 3 (720) | 6 (577) | Subhazard ratio; 2.21<br>(0.57 a 8.55); 0.250 |
| Time to invasive mechanical<br>ventilation in 14 days; n<br>events/person-time | 3 (382) | 6 (311) | Subhazard ratio; 2.30<br>(0.60 a 8.84); 0.230 |
| Time to invasive mechanical<br>ventilation in 28 days; n<br>events/person-time | 3 (718) | 6 (577) | Subhazard ratio; 2.19<br>(0.57 a 8.51); 0.260 |
| Time to death in 56 days; n events<br>/ person days | 4 (1473) | 8 (1339) | Hazard ratio; 2.56<br>(0.72 a 9.08); 0.147 |
| **Adverse events; No. events (%)** |  |  |  |
| Any event | 6 (19) | 12 (39) | Odds ratio; 2.74<br>(0.90 a 9.10); 0.085 |
| Serious event | 2 (6.2) | 6 (19) | Odds ratio; 3.60<br>(0.75 a 26.1); 0.14 |
| Infusion related event | 0 | 0 | NA |

NA: Not apply

**Fig 2.**
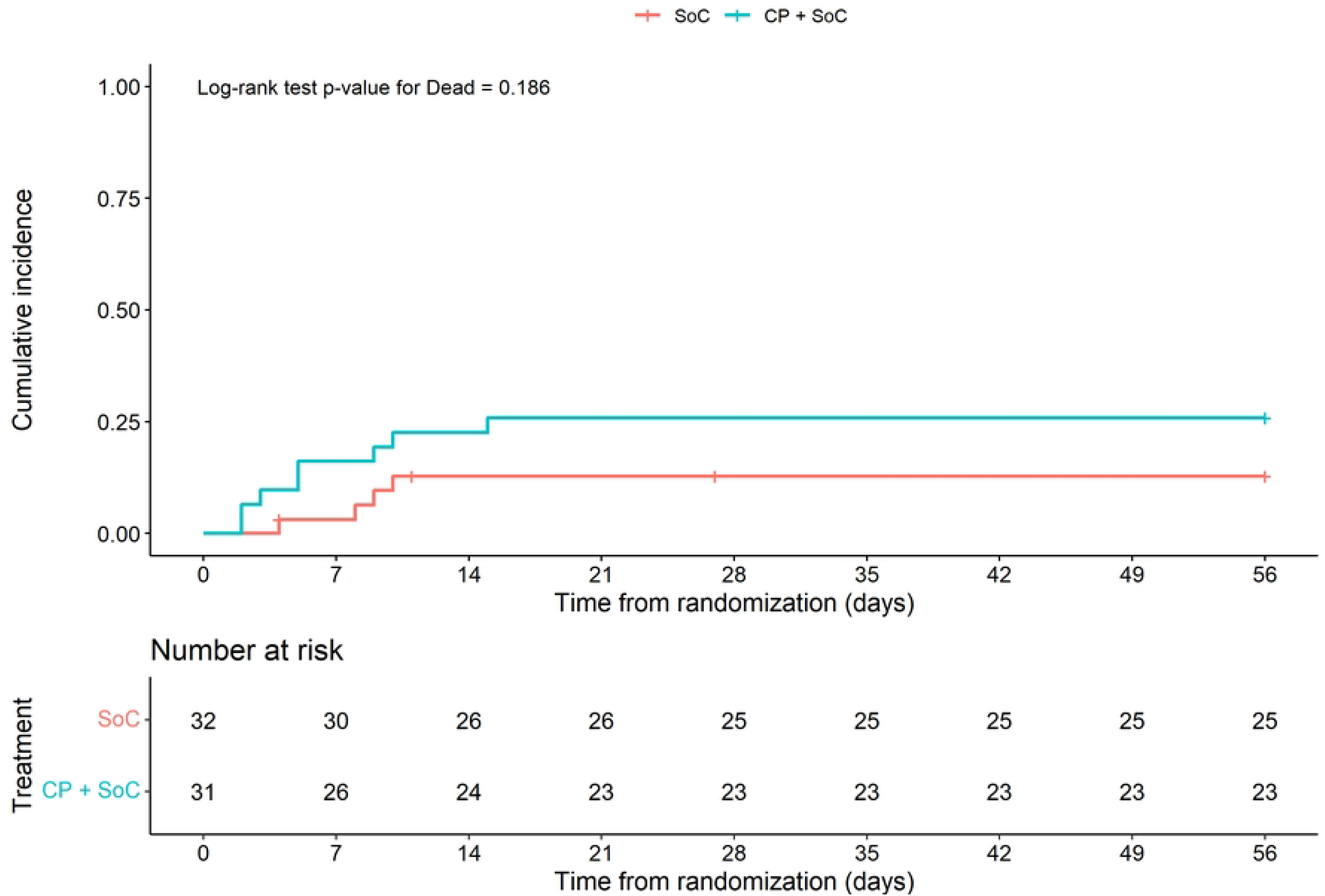
Inverse Kaplan-Meier curves for cumulative incidence of death after treatment with CP + SoC versus SoC alone

### Secondary efficacy outcomes

No statistically significant differences were found in the cumulative incidence curves for admission to the ICU within 28 days (p = 0.251) (Fig 3A). The incidence rate of admission to the ICU within 28 days was 10.4 per 1000 patient days in the CP + SoC group and 4.17 per 1000 patient days in the group that received only SoC. Considering death as a competitive event, the Fine and Gray model revealed no statistically significant differences in the incidence of ICU admission between both groups (subHR 2.06; 95% CI 0.57 to 8.55; p = 0.250). Compared to standard treatment alone, the estimated effect of convalescent plasma + standard treatment was the same for ICU admission at 14 days (subHR 2.21; 95% CI 0.57 to 8.55; p = 0.250).

**Fig 3.**
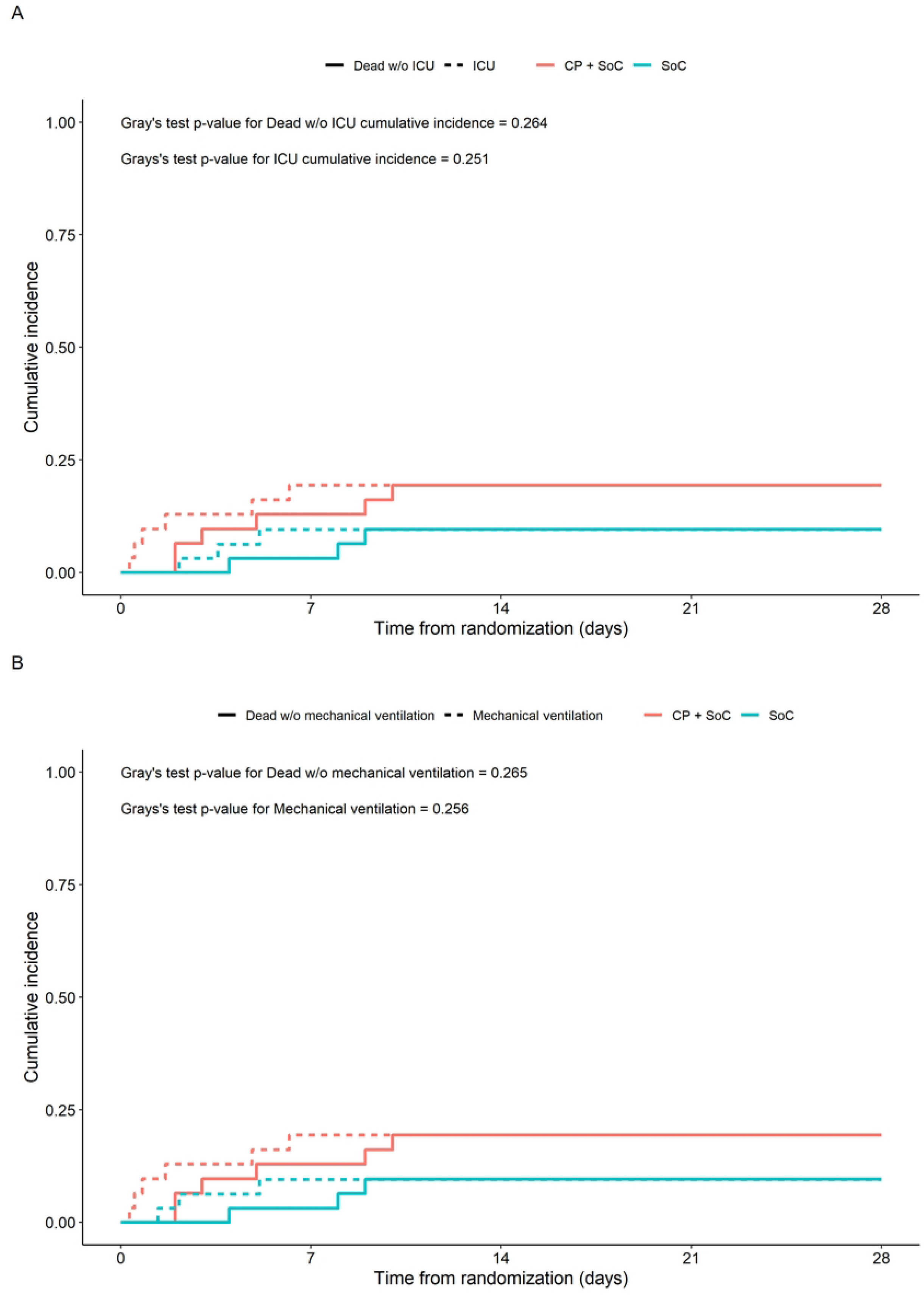
Cumulative incidence function curves for death (competing event) and (A) ICU admission or (B) mechanical ventilator admission after treatment with CP + SoC versus SoC alone

No statistically significant differences were found in the cumulative incidence curves for admission to mechanical ventilation at 28 days (p = 0.256) (Fig 3B). The 28-day incidence rate of invasive mechanical ventilation was 10.4 per 1,000 patient days in the convalescent plasma plus standard therapy group and 4.18 per 1,000 patient days in the standard therapy only group. Compared to standard treatment alone, the estimated effect of convalescent plasma + standard treatment was the same for admission to mechanical ventilation at 28 days (subHR 2.19; 95% CI 0.57 to 8.51; p = 0.260).

### Safety results

No infusion-related adverse events were reported in study participants. Adverse events were more common in the CP + SoC group (39%; 12 of 31 patients) than in the SoC group (19%; 6 of 32 patients). Similarly, serious adverse events were slightly more common in the CP + SoC group (19%; 6 of 31 patients) than in the SoC group (6.2%; 2 of 32 patients). However, there is high uncertainty regards the differences in the incidence of adverse events (OR 2.74; 95% CI, 0.90 to 9.10; p = 0.085) or serious adverse events (OR 3.60; 95% CI 0.75 to 26.1; p = 0.75) (Table 2 and S1 Table) if we consider the precision of these estimates and statistical significance.

## Discussion

This study aimed to assess the efficacy and safety of convalescent plasma (CP) plus standard of care (SoC) versus SoC alone in adult patients hospitalized with COVID-19 but not yet requiring mechanical ventilation. Our results found no evidence that PC had an effect in reducing mortality at 28 days. We also found no evidence that the frequency of adverse events was higher in those who received PC than those who received SoC.

Our results agree with those widely reported in the literature. Although initially, systematic reviews with meta-analyses (24-26) found evidence of benefit in favor of PC to reduce mortality, these included observational studies (27–30) and clinical trials with significant limitations (18,31). More recent clinical trials reported no evidence of the benefit of PC in reducing mortality, admission to the ICU, or mechanical ventilation (17,32-45). Later meta-analyses also concluded no evidence of PC efficacy in reducing the incidence of these outcomes (8,24,25,38,46–52). Clinical practice guidelines recommend against using PC in hospitalized patients with COVID-19 with a strong level of recommendation and a high certainty of evidence (14,15,53).

The RECOVERY (34), CONCOR-1 (44), and REMAP-CAP (45) studies were the three largest clinical trials conducted to assess the efficacy and safety of convalescent plasma, and none found evidence of a benefit of high-dose CP in reducing mortality, ICU admission or mechanical ventilation in patients with COVID-19. Like our study, all of them were open-label and were stopped early. The RECOVERY trial (34) enrolled 11,558 patients (5,795 received CP + SoC and 5,763 received SoC). The study found evidence in favor of no significant differences (RR = 1.00; 95% CI 0.93-1.07) in 28-day mortality and other hospital outcomes such as mechanical ventilation. The CONCOR-1 trial (44), which enrolled 614 patients in the CP group and 307 in the SoC group, found no significant difference in its primary outcome of intubation or death at day 30 (RR = 1.16; 95%CI 0.94-1.43) nor in its secondary outcomes such as mortality, admission to intensive care and hospital stay. The REMAP-CAP trial (45), which enrolled 1084 critically ill patients in the PC group, and 916 in the control group, found no significant differences in in-hospital mortality outcomes. However, it did report potential for harm in patients who received convalescent plasma after the seven days of hospitalization.

Regarding the safety of PC, to date, 51 clinical trials have been published that evaluated the use of PC, concluding, through a meta-analysis, that with a low degree of certainty, PC does not increase the occurrence of adverse events (15). Consistent with existing evidence, our study did not find any transfusion-related SAEs and, although there was a higher frequency of adverse events of any kind in the group treated with PC + SoC compared to the SoC group, these differences were not statistically significant.

Observational surveillance studies suggest that adverse reactions are infrequent and related to conventional risks of plasma infusion for other indications. For example, a study evaluating safety using records from 5,000 clinicians of hospitalized adult patients with severe COVID-19 found a low mortality rate of 0.3%. Likewise, the incidence of all serious adverse events (SAEs) in the first four hours after the transfusion was less than 1% (54). In addition to death (4 cases of 25 related SAEs), the main SAEs were transfusion-related circulatory overload (7 of 25 related SAEs), transfusion-related acute lung injury (11 of 25 SAEs), and severe transfusion-related allergic reactions (3 of 25 EAS). Months later, the update of this study extended the analysis to 20,000 patients, confirming the low frequency of adverse events: <1% for thrombotic and thromboembolic events and ∼3% for cardiac events (55).

This study has limitations to be considered. All patients had moderate to severe COVID-19, so our conclusions cannot be extrapolated to other groups of patients with different degrees of severity, especially patients with mild COVID-19. Another limitation is that the trial was open label, which could have influenced more subjective outcomes such as the recognition and/or reporting of some adverse events. However, these results are unlikely to have influenced hard outcomes such as mortality, ICU admission, or admission to mechanical ventilation.

In conclusion, in our study, using CP + SoC in patients with moderate COVID-19 did not reduce mortality or improve other clinical outcomes at day 28 compared to SoC alone. Our results are consistent with the literature on the lack of benefit of CP and reinforce the evidence in favor of discouraging CP use in hospitalized patients with moderate to severe COVID-19.

## Data Availability

The raw data was generated at the Institute for the Evaluation of Technologies in Health and Research - IETSI of Peru's Social Security of Health (EsSalud). Restrictions apply to the public availability of these data due to institutional patient data sharing policies. However, the data is available upon reasonable request from the author.

